# Real-world Safety and Efficacy of Dimethyl Fumarate in Relapse-Remitting Multiple Sclerosis Patients: A Regional Cohort Report of the Iranian Patients

**DOI:** 10.64898/2026.07.31.26359413

**Authors:** Masoud Etemadifar, Fateme Jannesari, Mohsen Raeisidehkordi, Kamran Rezaei, Mehri Salari, Mahdi Norouzi

**Author notes:** Corresponding Author: **Email:**, **Address:** Hezar Jerib Street, Isfahan, Iran 8174673461.

## Abstract

**Background:** Real-world evidence evaluating the long-term effectiveness and safety of dimethyl fumarate (DMF) in relapsing-remitting multiple sclerosis (RRMS) remains limited, particularly in Middle Eastern populations. Furthermore, whether previous exposure to disease-modifying therapies influences longitudinal treatment response has not been adequately characterized. We evaluated the real-world effectiveness, safety, and temporal treatment dynamics of DMF in RRMS and compared outcomes between treatment-naïve and previously treated patients.

**Methods:** This longitudinal observational cohort study enrolled 120 adults with RRMS initiating DMF (TECRA®) at two multiple sclerosis centers in Iran. Clinical outcomes, magnetic resonance imaging (MRI) activity, disability progression, and adverse events were assessed over 18 months at 6-month intervals. Repeated Expanded Disability Status Scale (EDSS) measurements were analyzed using linear mixed-effects models, while relapse counts and MRI lesion activity were evaluated using generalized estimating equations. Prespecified subgroup analyses examined differences according to prior treatment status.

**Results:** Ninety-five patients completed the study. DMF produced a marked suppression of disease activity, reducing the annualized relapse rate by 95% (1.56 ± 0.93 to 0.08 ± 0.24; *P* < 0.001). EDSS improved during the first year and remained near baseline after 18 months despite a modest increase during the final follow-up interval. MRI inflammatory activity declined significantly throughout follow-up, although a mild increase in gadolinium-enhancing lesions after 12 months suggested possible attenuation of treatment effect over time. Overall, 88.4% of patients remained relapse-free, 70.5% demonstrated no MRI disease activity, and 64.2% achieved no evidence of disease activity (NEDA-3). While overall clinical outcomes were comparable between treatment-naïve and previously treated patients, longitudinal analyses revealed distinct temporal patterns of MRI activity between groups. DMF was well tolerated, with predominantly mild cutaneous and gastrointestinal adverse events and infrequent treatment discontinuation.

**Conclusions:** In conclusion, DMF was well tolerated and effective in reducing clinical and radiological disease activity. These findings support the long-term effectiveness of DMF in routine clinical practice while highlighting the importance of continued clinical and radiological monitoring to optimize individualized treatment strategies.

## Introduction

Multiple sclerosis (MS), characterized by demyelination, gliosis, and axonal loss, is the most common chronic autoimmune disease of the central nervous system, mostly presenting as relapsing-remitting MS (RRMS) (1). RRMS, compared to other MS subgroups, has a benign early course. However, left uncontrolled, the cumulative axonal damage from recurrent relapses may lead to progressive disability. In terms of controlling the disease, more than a dozen disease-modifying treatments (DMTs) are available, ranging from injectable medication (Interferon β, Glatiramer acetate, etc.) to oral medications (Dimethyl fumarate (DMF), Teriflunomide, Fingolimod, Siponimod, etc.) and, more recently, the monoclonal antibodies (Natalizumab, Alemtuzumab, Ocrelizumab, etc.) (1).

Dimethyl fumarate (DMF) approved in 2013 for relapsing MS is an oral ester of fumaric acid. DMF is widely in use as the first line DMT in RRMS due to its favorable efficacy–safety balance (2). It exerts widespread anti-inflammatory and cytoprotective effects. Mechanistically, monomethyl fumarate (active metabolite of DMF) activates the Nrf2 (NF-E2–related factor 2) antioxidant pathway, leading to upregulation of antioxidant and phase II detoxifying genes (e.g., NQO1, HO-1) (3). DMF modulates immune responses through decreased pro-inflammatory cytokine production, lymphocyte migration, and increased regulatory cell subsets. Landmark phase III trials (DEFINE, CONFIRM) have indicated the pronounced DMF properties in controlling and deescalating the clinical and radiological disease activity features in RRMS patients (4,5). Same results have been shown in long-term extension trials (e.g., ENDORSE) (6). This study aims to assess the safety and efficacy properties of *DMF*. Furthermore, the effect of DMF within the treatment-naive and pre-treated patients has rarely been investigated.

## Materials and Methods

### Participants and Study Design

This study is a longitudinal observational cohort study in two MS centers in Isfahan, Iran. We screened over 1000 patients, of whom we enrolled 120 patients with RRMS undergoing TECRA® therapy. All participants provided written informed consent, and the study protocol was approved by the local ethics committee (approval code IR.MUI.MED.REC.1402.170) in accordance with the Declaration of Helsinki. The inclusion criteria were as follows:

- Written informed consent provided.
- Definite RRMS diagnosis based on the 2017 McDonald criteria.
- Age between 18 and 50 years.
- Initiation of DMF treatment during the study period.
- No other concurrent immunomodulatory treatments.
- Availability of essential baseline and follow-up data in the patient’s medical records.

### Treatment Protocol and Outcome Measures

Our primary outcome was assessing the efficacy-safety balance of DMF (TECRA®, Alborz Zagros Pharmaceutical Co., Iran) in RRMS patients. Based on the protocol, patients were started on daily DMF 120 mg for the first week, followed by 120 mg BD for another four weeks. Following this, a dosage of 240 mg BD capsule was taken by the patients for the rest of the treatment duration. The disease endpoints were captured in three categories: (a) clinical data, (b) MRI findings, and (c) adverse events and discontinuation. Based on the successive visits and the patient’s dossier, clinical and MRI data were collected retrospectively for the 6 months preceding DMF initiation (baseline period) and prospectively for 18 months following treatment initiation. The secondary outcome was set as the treatment efficacy and safety profile in treatment switchers and treatment-naïve patients. The analysis intervals were set equal to the pretreatment interval: 6 months before initiation, 0-6 months, 6-12 months, and 12-18 months.

Relapses, Expanded Disability Status Scale (EDSS) scores, and MRI outcomes (new/enlarging T2 lesion count and number of gadolinium-enhancing lesions) were recorded at regular intervals. A relapse was defined as a new or worsening neurological symptom lasting> 24 h without other strong explanations. Subsequently, the annualized relapse rate (ARR) was calculated by dividing relapse counts by years. Confirmed disease progression (CDP) was defined as an EDSS increase sustained for 6 months:

≥1.0 point (if baseline EDSS is ≤5.5)
≥0.5 point (if baseline EDSS is >5.5)

Likewise, the confirmed disease improvement (CDI) was defined as a ≥1.0-point decrease in EDSS from baseline, sustained for 6 months. Furthermore, the no evidence of disease activity (NEDA-3) was computed, which is a composite score of patients with: (a) no relapses, (b) no new/enlarging T2 or gadolinium-enhancing lesions on MRI, and (c) no CDP. Adverse events (AEs) and discontinuation rates, and reasons were also gathered.

### Statistical Analysis

Analyses were performed in SPSS version 27 (IBM Corp., Armonk, NY). A per-protocol approach was used in interpreting the treatment outcomes. We treated the longitudinal data in each interval as repeated measures. For continuous outcomes (EDSS scores), we used a linear mixed effects model with patient as a random effect and treatment group and time as fixed effects. Relapses and MRI lesions are count data; so, they were analyzed using generalized estimating equations (GEE) with a Poisson distribution and log link. The Wilcoxon rank-sum test was used for non-repeated continuous variables. All statistical tests were two sided with a significance level of 0.05. Continuous data are presented as mean ± standard deviation (SD), and categorical data as proportions.

## Results

### Baseline Characteristics

Demographic and baseline characteristics of included patients are presented in Table 1. Of 120 included patients, 95 patients completed 18-month follow-up per protocol (22 were lost to follow-up, and 3 discontinued DMF). The patient attrition during follow-up was mostly due to untraceable patients who missed their routine visits. The mean age of the patients was 36.03 ± 7.82, with a strong female preponderance (77%). Disease duration was almost 8 ± 7 years, with a baseline EDSS of 1.35 ± 0.55 throughout the whole cohort. The baseline ARR of the included patients was 1.56 ± 0.93. Thirty-eight patients were previously treated with other DMTs, while 57 patients were treatment naïve. Apparently, the outcomes, except for EDSS, due to its cumulative nature, were more pronounced in treatment-naïve patients.

**Table 1.**
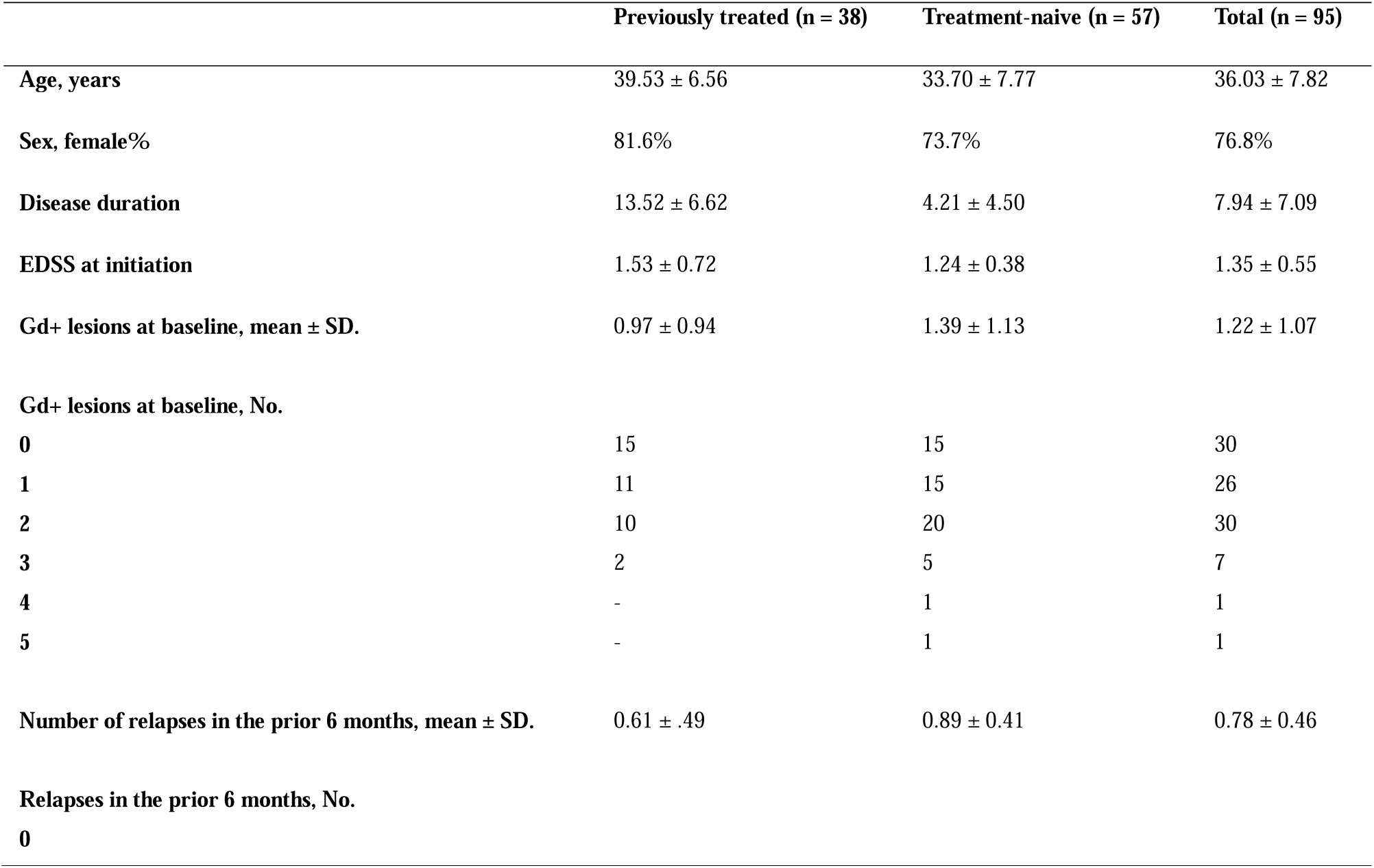
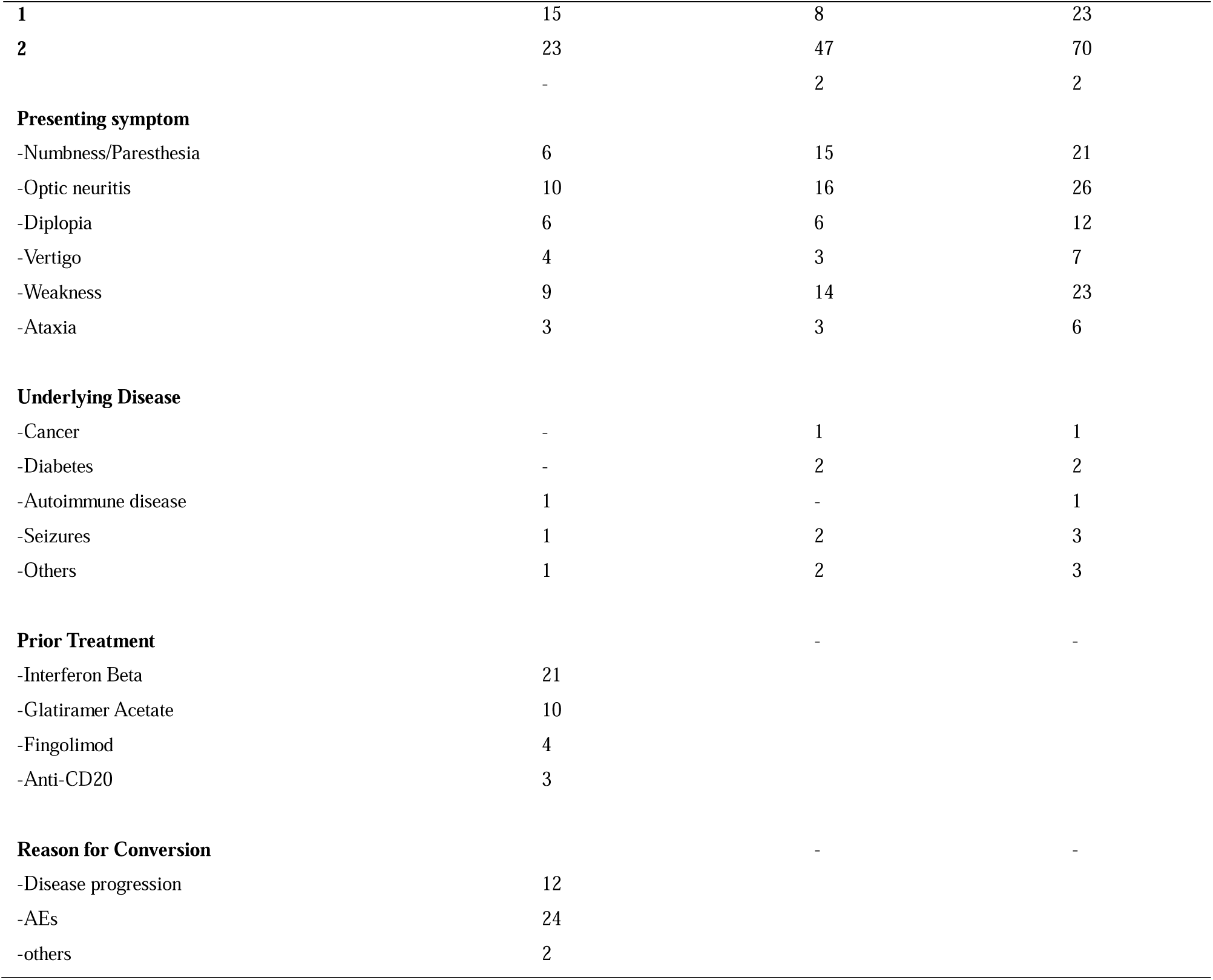
Baseline characteristics of the patients.

### Treatment Outcomes

Longitudinal analysis of repeated EDSS at four timepoints, with 6-month intervals, indicated that the overall changes of EDSS through the follow-up duration were significant (Figure 1). Mean EDSS decreased significantly in the first 6 months (by 0.08 points, P = 0.01) and stayed lower than baseline at 12 months. By 18 months, EDSS had crept back up slightly, but remained close to the baseline level (no significant difference from baseline (MD =0.063, P = 0.070) or 12 months (MD = −0.032, P > 0.05)). Additionally, leaving out the baseline EDSS, post-treatment analysis indicated a significant change in EDSS post-treatment, with stability in the first two intervals and a slight increase in the third one.

**Figure 1.**
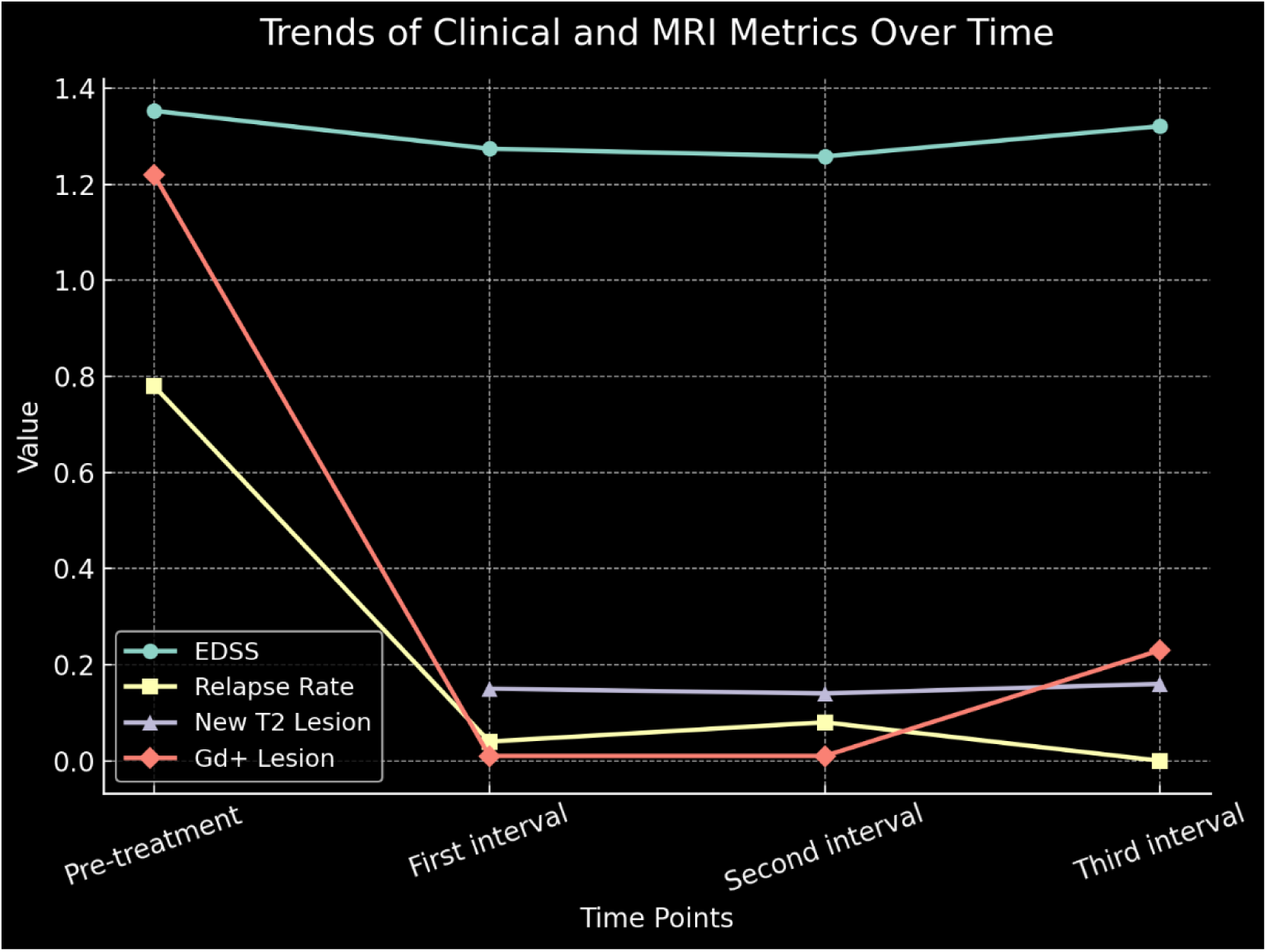
Whole-cohort clinical and MRI outcomes over time, shown at 6-month intervals (1: baseline; 2: 0–6 months; 3: 6–12 months; 4: 12–18 months).

Post-treatment ARR fell by 95% with DMF treatment (from 1.56 ± 0.93 to 0.08 ± 0.24 (p< .001)). Likewise, a significant reduction in relapse rate was indicated during the follow-up duration (P<0.001). Relapses in all three post-treatment intervals were significantly lower than baseline. Notably, the relapse rate didn’t increase in the second year; contrary to EDSS, there was even a lower rate in the months 12-18 compared to 6-12 (P = 0.019). 88.4% of the patients remained relapse-free.

Regarding the Gd-enhanced lesions, the number of lesions reduced significantly over time (P<0.001). Each post-treatment interval indicated significantly fewer Gd-enhancing lesions compared to baseline. However, MRI inflammatory activity remained significantly lower than baseline throughout follow-up. A modest increase in gadolinium-enhancing lesions was observed during the 12–18-month interval compared with earlier follow-up periods. During the follow-up, 88.4% of patients were relapse-free. MRI was clear in 70.5% of the patients (Gd+ lesion-free in 85.3% and new T2 lesion-free in 74.7%). 2.1% of patients indicated CDI, while none indicated CDP. Overall, 64.2% of the patients achieved no signs of disease activity (NEDA-3).

### Adverse Events and Tolerability

The treatment persistence rate after 18 months among 120 enrolled patients was 79.2%. Of the three patients who discontinued the treatment, one patient refrained from the treatment due to adverse events (gastrointestinal). The treatment was ceased in two patients due to inefficacy. The patients who discontinued the treatment were from the pre-treated group. 36% of patient experienced AEs (Table 2). Skin and cutaneous manifestations were among the most common AEs (24.2%). These manifestations were more prominent in the treatment-naïve group compared to the treated group. On the other hand, the gastrointestinal AEs were more prominent in the pre-treated group. Other less common AEs included respiratory manifestations (2.2%), fatigue (1.1%), headache (4.2%), and sleep disorders (1.1%).

**Table 2.**
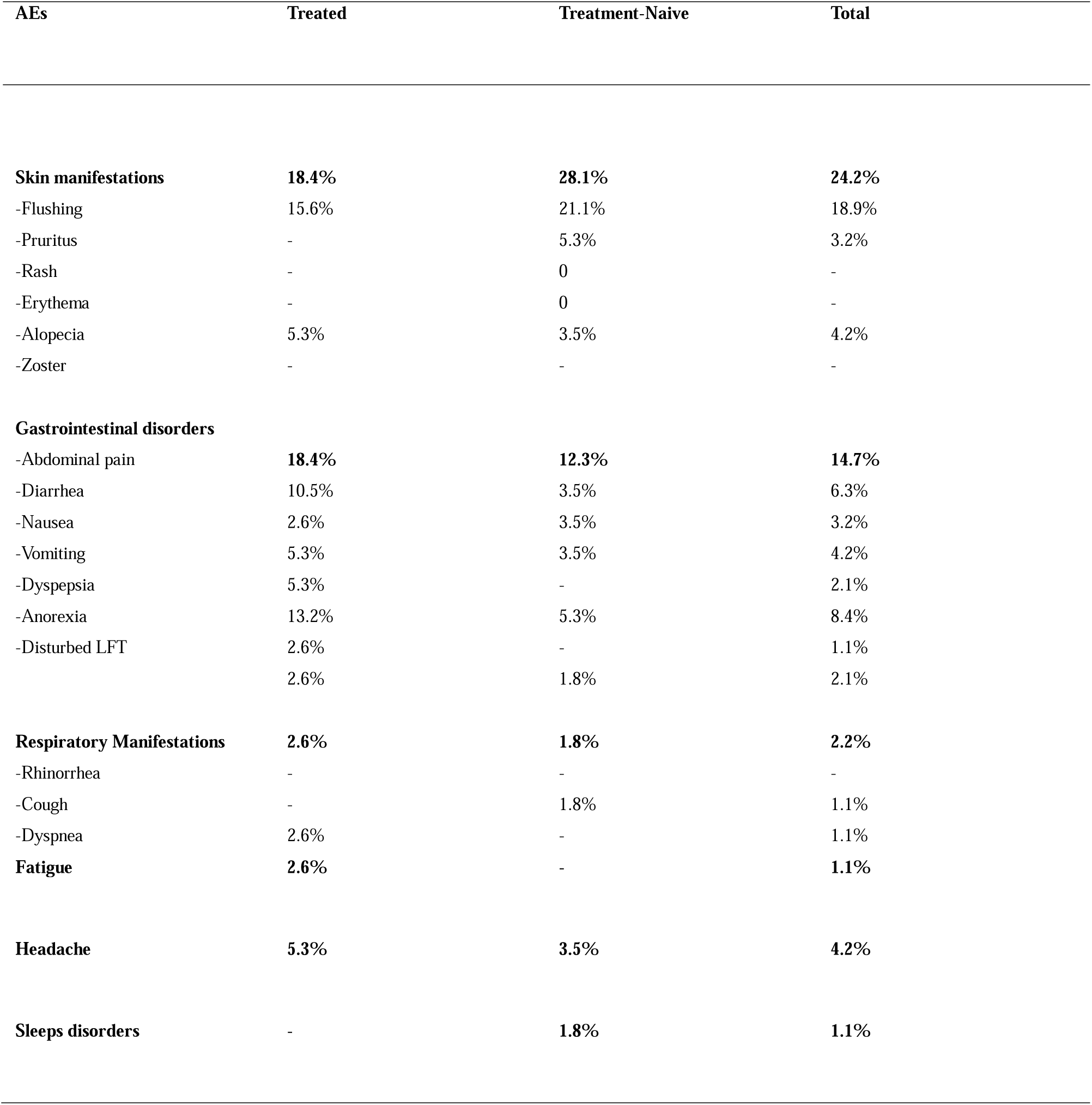
Main adverse events in patients undergoing *DMF*.

| AEs | Treated | Treatment-Naive | Total |
| --- | --- | --- | --- |
| **Skin manifestations** | **18.4%** | **28.1%** | **24.2%** |
| -Flushing | 15.6% | 21.1% | 18.9% |
| -Pruritus | - | 5.3% | 3.2% |
| -Rash | - | 0 | - |
| -Erythema | - | 0 | - |
| -Alopecia | 5.3% | 3.5% | 4.2% |
| -Zoster | - | - | - |
| **Gastrointestinal disorders** |  |  |  |
| -Abdominal pain | **18.4%** | **12.3%** | **14.7%** |
| -Diarrhea | 10.5% | 3.5% | 6.3% |
| -Nausea | 2.6% | 3.5% | 3.2% |
| -Vomiting | 5.3% | 3.5% | 4.2% |
| -Dyspepsia | 5.3% | - | 2.1% |
| -Anorexia | 13.2% | 5.3% | 8.4% |
| -Disturbed LFT | 2.6% | - | 1.1% |
|  | 2.6% | 1.8% | 2.1% |
| **Respiratory Manifestations** | **2.6%** | **1.8%** | **2.2%** |
| -Rhinorrhea | - | - | - |
| -Cough | - | 1.8% | 1.1% |
| -Dyspnea | 2.6% | - | 1.1% |
| **Fatigue** | **2.6%** | - | **1.1%** |
| **Headache** | **5.3%** | **3.5%** | **4.2%** |
| **Sleeps disorders** | - | **1.8%** | **1.1%** |

### Subgroup Analysis

By the end of 18 months, there were no significant differences between treatment-naïve and previously treated patients in terms of relapse count, EDSS change, or MRI lesion counts (p > 0.05 for all comparisons). However, longitudinal analyses revealed some differences in the pattern of MRI outcomes over time between the two groups. An interaction effect between time and prior treatment status was observed for new T2 lesions: treatment-naïve patients had more new T2 lesions in the first 6 months, but fewer in the subsequent 12 months, compared to the previously treated group (interaction p = 0.045). A similar subtle interaction was seen for Gd+ lesions (interaction p = 0.048), with treatment-naïve patients overall exhibiting a higher rate of enhancing lesions, but with different temporal dynamics. In contrast, no significant group differences or interactions were found for relapse rate or EDSS change over time (Figures 2 and 3).

**Figure 2.**
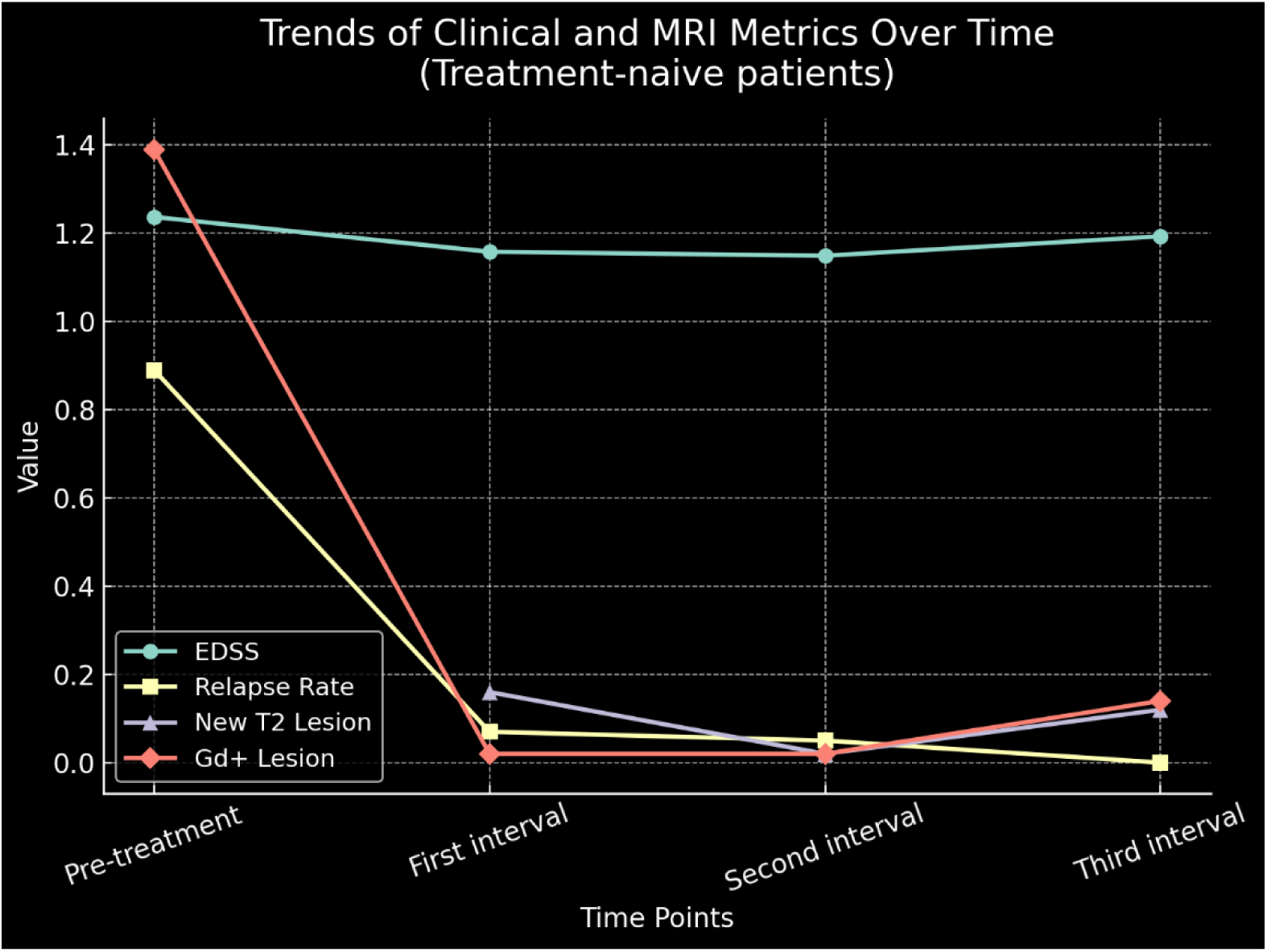
Longitudinal clinical and MRI outcomes in treatment-naïve patients (intervals as defined in Figure 1).

**Figure 3.**
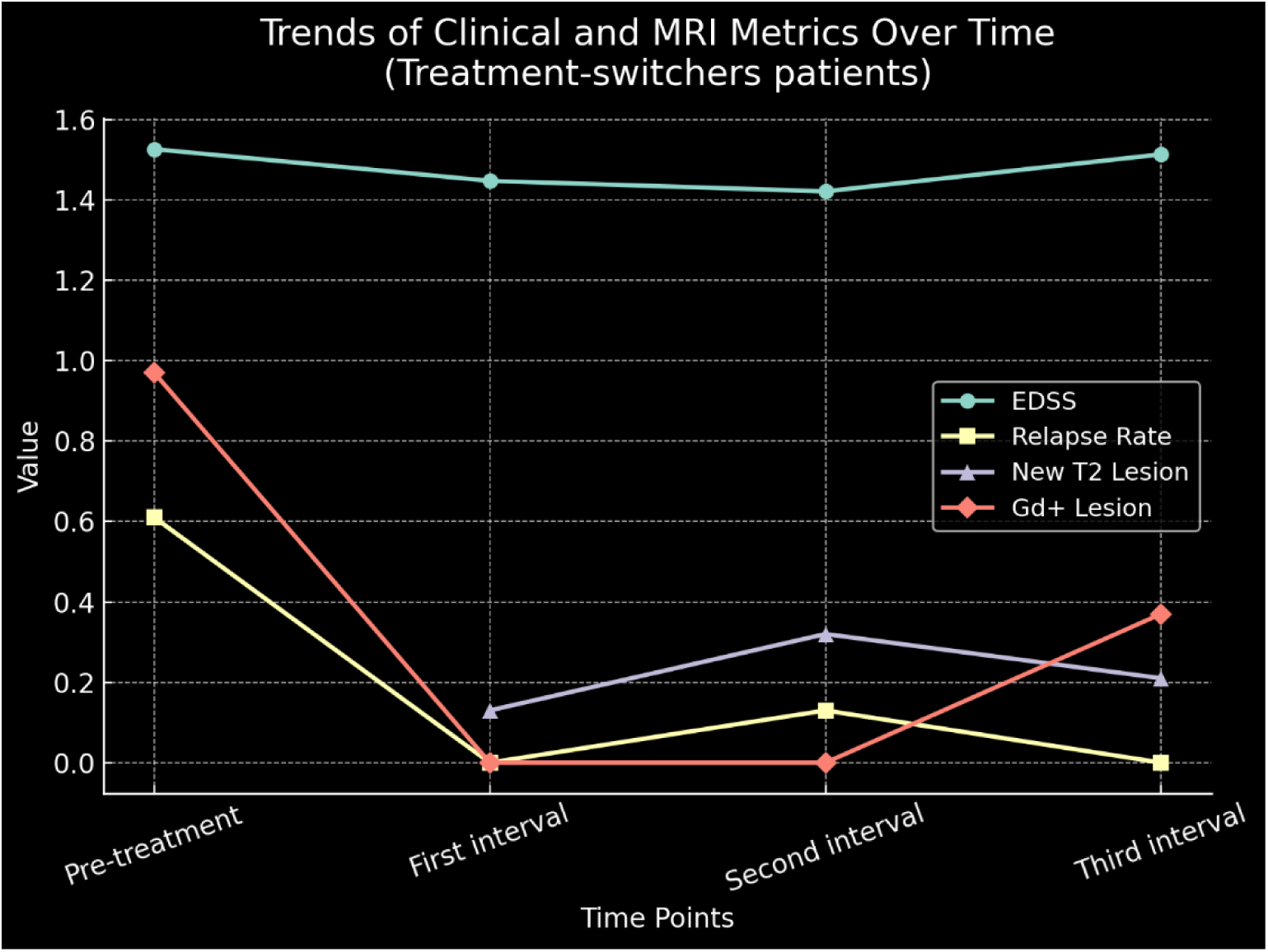
Longitudinal clinical and MRI outcomes in previously treated patients (intervals as defined in Figure 1).

## Discussion

This prospective cohort study highlights the DMF efficacy in suppression of disease activity in RRMS in both pre-treated and treatment-naïve patients. Over 18 months, we observed significant reductions in annualized relapse rates, suppression of disability progression, and MRI features of disease activity. These findings align with pivotal trials—DEFINE and CONFIRM—which reported a 44–53% reduction in relapse and a 71–90% decrease in Gd+ lesions compared to placebo at 2 years (5,7). Similarly, the ENDORSE extension study showed sustained low ARR (≈ 0.15) over a span of 10 years (6). Early disease activity likely contributes to escalated long-term risk of disease worsening and disability, along with increasing the risk of conversion to secondary progressive MS (8–10). Fortunately, the early suppression of disease activity with DMF is encouraging.

Despite this robust initial efficacy, we observed a mild resurgence of disease activity in the second year of treatment. By around 12 months, there was a slight increase in mean EDSS and a rise in new Gd+ lesions. Notably, no patient met the formal criteria for CDP at 18 months, although this zero CDP should be interpreted cautiously because EDSS measurements require 6-month confirmation (the final EDSS could not be confirmed due to insufficient follow-up time). From an immunological perspective, early DMF-induced changes in regulatory T cells and cytokine profiles, along with gradual lymphocyte reconstitution, may underlie this “escape” phenomenon (3). In practical terms, medication adherence might decline over time, though reported persistence was high in our cohort (which could be an overestimate). DMF’s known side effects (e.g., gastrointestinal upset and flushing) can impact long-term compliance. Another possibility is subclinical progression independent of relapses (PIRA) contributing to the slight EDSS increase, although we did not assess PIRA in this study. Whether this observation reflects natural variability, changes in disease biology, or treatment-related dynamics remains uncertain and warrants investigation in larger cohorts with longer follow-up. Additionally, these borderline effects should not be treated as significant divergence in response to the treatment. The differences are far from clinical relevance. We have reported them just as an exploratory observation. A longer follow-up duration or a larger cohort may fade these borderline differences.

Differences in baseline disease activity between the two groups (treatment-naïve vs. previously treated) may have influenced our findings. The treatment-naïve patients had higher inflammatory activity at baseline (mean Gd+ lesions 1.39 vs 0.97), consistent with earlier-stage MS (11). Whereas previously treated patients had a greater chronic lesion burden (more T2 lesions). Both groups showed significant reductions in gadolinium-enhancing lesions on therapy; however, the previously treated group exhibited a modest increase in enhancing lesions during the 12–18 month interval. All patients received the same DMF dosing regimen, raising the question of whether those with very active disease (especially those previously treated with high lesion burdens) might benefit from an earlier switch to more potent therapies (12).

Several limitations affected our findings. The observational design and patient attrition during follow-up may have introduced selection bias despite the use of longitudinal statistical methods. Although treatment persistence was high, missing follow-up data may have influenced estimates of long-term treatment effects. We intended to analyze lymphocyte count trends, but this was not feasible due to the large amount of missing data. Furthermore, assessing different dosages and regimens of DMF to reach optimum safety-efficacy in both groups is a direction for future studies. Additionally, assessing the PIRA existence and progression, and the clinical significance of it, should be considered by future studies.

## Conclusion

In summary, DMF is effective at reducing relapses and MRI lesion activity in RRMS. DMF demonstrated sustained reductions in clinical and radiological disease activity over 18 months with a favorable safety profile. Continued clinical and MRI surveillance remains important to characterize long-term treatment response and support individualized therapeutic decision-making. Differences between naïve and previously treated patients likely reflect differences in the baseline features. These findings underscore the importance of a personalized treatment plan and ongoing vigilance.

## Data availability

The datasets generated and/or analyzed during the current study are available from the corresponding author on reasonable request.

## Author contributions

M.E. and M.N. conceived and designed the study. M.N. contributed to manuscript writing, statistical analysis, and reviewing the manuscript. M.R., M.E, and F.J. contributed to data curation. K.R. and M.S. contributed to manuscript review. All authors have read and approved the final version.

## Acknowledgement

We have used ChatGPT-4o to enhance the readability of the manuscript.

